# Drone-delivered Automated External Defibrillators for out-of-hospital cardiac arrest. A simulation study

**DOI:** 10.1101/2024.02.23.24303253

**Authors:** Christopher M Smith, Joe Phillips, Nigel Rees, Carl Powell, Anthony Sheehan, Mary O’ Sullivan

## Abstract

**Background:** Cardiopulmonary resuscitation (CPR) and defibrillation at least doubles survival to hospital discharge following out-of-hospital cardiac arrest. Members of the public can perform both before the ambulance service arrives. However, bystanders use a public-access Automated External Defibrillator (AED) in around 5% of cases. Using Unmanned Aerial Vehicles (‘drones’) to deliver AEDs may overcome many of the barriers preventing public-access AED use.

We investigated how quickly and easily bystanders performing CPR could use drone-delivered AEDs.

**Methods:** We developed an AED-capable drone between May and November 2020. In July and September 2021, we conducted eighteen out-of-hospital cardiac arrest simulations. A single participant found a simulated patient inside a building and made a 999-call to a Welsh Ambulance Services NHS Trust call-handler. Once cardiac arrest was confirmed during the 999-call a nearby drone launched, reached hovering altitude and delivered the AED immediately outside the building. The participant retrieved the AED when instructed to do so, attached it to the patient and delivered a single shock.

The primary outcome was hands-off CPR time. We investigated ease of AED retrieval via a questionnaire adapted from the System Usability Scale and explored participant behaviours via brief post-simulation interviews and reviews of audio (999-call) and video recordings of the simulation.

**Results:** Hands-off CPR time was (median) 109s (interquartile range 87-130s). Participants spent 19s (16-22s) away from the patient’s side when retrieving the AED. They found it easy to use the AED but often sought reassurance from the call-handler that it was appropriate for them to retrieve it.

**Conclusion:** Participants found it easy to retrieve and use an AED delivered by drone in simulated out-of-hospital cardiac arrests. Hands-off CPR time was potentially clinically relevant in this lone bystander simulation, but there was only a small increase in hands-off time caused by retrieval of the drone-delivered AED.

What is already known on this topic – summarise the state of scientific knowledge on this subject before you did your study and why this study needed to be done
Drones have been used to deliver AEDs in simulation studies across the world and in a real-life pilot in Sweden. Real-world success is so far limited, and no functioning system for this to happen in real-world out-of-hospital cardiac arrests in the UK.

What this study adds – summarise what we now know as a result of this study that we did not know before
We have demonstrated a feasible drone-delivered AED system. Lone bystanders spent a median of 19 seconds away from the patient to retrieve the drone-delivered AED. Interaction with the drone and AED was not difficult, and the 999 call-handler has a vital role in facilitating safe and timely retrieval of the drone-delivered AED.

How this study might affect research, practice or policy – summarise the implications of this study
Lone bystanders are currently not instructed by ambulance services to leave a patient to retrieve a nearby public-access AED, but collecting an AED delivered directly to them may be appropriate.
The next step in developing a drone-delivered AED system for real-world use in the UK is to integrate a drone-delivered AED system into an ambulance service’s Emergency Operations Centre system and to test the out-of-hospital cardiac arrests response in prolonged ‘beyond visual-line-of-sight’ drone flights.

## Introduction

Around 10% patients who sustain an out-of-hospital cardiac arrest (OHCA) survive to hospital discharge (1-3). Good quality cardiopulmonary resuscitation (CPR) and prompt defibrillation using an Automated External Defibrillator (AED) substantially increase survival rates (4).

Bystander use of public-access Automated External Defibrillators (AEDs) before the arrival of the ambulance service is associated with increased survival to hospital discharge (OR 1.73) and survival with good neurological function (OR 2.12) (5). However, in the UK (2020), an AED was used by a member of the public in only 4.4% of cases (1).

Improving bystander AED use to improve survival to hospital discharge is a key aim of resuscitation strategies from all four UK home nations (6-9). Many barriers to timely AED attachment relate to a bystander’s ability to promptly locate, access and retrieve a public-access AED (10).

One way of overcoming this may be to deliver AEDs by Unmanned Aerial Vehicles (UAVs, ‘drones’). A drone-delivered AED network is now operational in Sweden (11) and there was a recent case report of survival to hospital discharge in a patient defibrillated by a drone-delivered AED in this system (12). Other studies have modelled optimal drone location (13, 14) and flights for simulated OHCAs (15-18).

Our aim was to determine whether a lone bystander performing CPR on a simulated patient could effectively retrieve and attach a drone-delivered AED.

## Methods

### Developing the delivery system

We compared two systems to deliver an AED by drone in controlled, closed-test conditions.

a. The drone hovers over the delivery site (at a height of 7-10m) and lowers the AED by winch. Once the AED is on the ground, the winch is retracted and the drone hovers above the AED to indicate its location.
b. The drone lands on the ground at the delivery site, the AED detaches and the drone hovers above the AED to indicate its location

We conducted five test flights for both delivery mechanisms – so the drone operator developed sufficient experience – and recorded time for AED delivery. This started when the drone was at the delivery site and began to (a) extend its winch or (b) descend to the ground, and ended when the drone had returned to its hovering position. We considered that this endpoint was when it would be safe for a bystander to retrieve the AED.

TETRA Drones Ltd (https://www.tetradrones.co.uk/) developed the drone delivery systems, and it carried a Lifepak CR2 AED Training Unit (Physio Control).

### Simulations

We conducted OHCA simulations on 10/07/2021 and 04/09/2021 at the Sussex Police Training Centre – a controlled-access site with mocked-up buildings and roads. We placed a CPR manikin (Laerdal Medical) in a single-storey building from which one could get outside via a hallway and an external door. Immediately outside was a pavement and road.

We informed participants that: they would find a simulated patient in the building who was unconscious and not breathing; they should indicate when they wished to make an emergency (999) call and help the patient as they saw fit; at some point an AED would be delivered by drone; and they should retrieve it when we indicated it was safe to do so

A study team member observed the participant but did not intervene, other than to hand them a mobile phone with a pre-programmed number for the simulated 999-call. The participant was connected to a Welsh Ambulance Services NHS Trust (WAST) call-handler. They were operating at a training centre but handled it as if it were a ‘live’ 999-call, using the Advanced Medical Priority Dispatch System (AMPDS) to prioritise the call.

Once the call-handler recognised a potential OHCA there was an automatic allocation to a mobile device (Terrafix Ltd, http://www.terrafix.co.uk/) allocated to a study team member from WAST (CP). He was situated with the drone operator and then cued them to initiate flight. For these simulations the drone was situated out-of-sight of participants, approximately 50m away. It flew to the delivery site and delivered the AED.

The study team indicated when it was safe to approach the drone by sounding a loud siren. Additionally, the 999 call-handler gave the following instruction:

*“A defibrillator has been dispatched to your location by drone. When it has arrived, and it is safe to approach a horn will sound allowing you to retrieve it safely. Please let me know as soon as it is right next to the patient*.*”*

Simulations were timed: starting when participants entered the building and ending once participants resumed CPR after attaching the AED and delivering a shock. There were two fixed position video cameras: one inside directed at the manikin, and one outside directed at the drone delivery site. WAST made audio recordings of the 999-call on the first study day, but this did not happen (inadvertently) on the second day. The phone audio on the second study day was audible via the video recording. CMS made field notes based on the observations of the study team during simulations.

We asked participants to complete a short post-simulation questionnaire, based on the ‘System Usability Scale’ (SUS). This was developed in industry to assess the usability of a new device or system and has been made freely available for usability assessments (19). CMS also conducted a brief (<5min) interview asking participants about their experience of retrieving the AED. Interviews were based on the ‘Learning Conversation’ used in resuscitation courses to evaluate behaviours during mock cardiac arrest simulations (20).

The primary outcome was ‘hands-off CPR time’ – the time between stopping chest compressions to retrieve the AED to starting chest compressions again after the AED delivered its shock. Secondary outcomes were: time spent away from patient retrieving AED; time from start of 999 call to first shock; SUS scores; findings from post-simulation interview; findings from video and audio call analyses.

We have reported study timings and post-simulation questionnaire scores (1:strongly disagree–5:strongly agree) as median (interquartile range, IQR). JP directly observed participants in simulations and made notes between simulations. CMS performed post-simulation interviews, making notes at the time. CMS typed up these contemporaneous handwritten notes and added further information after reviewing audio and video of the simulations.

CMS and JP reviewed the summated information to identify key themes relating to the participants’ interaction with the AED, drone and 999-call-handler. They then added their reflections, independently considered important issues and grouped them into relevant themes. They decided final themes by consensus.

We adopted an iterative approach to understanding and synthesising information to make it accessible to those reading, which is common to thematic analyses (21). We attempted to improve reliability of findings by triangulating data from multiple sources and organising it into written notes before analysis and presentation (22). We attempted to generate themes inductively with little pre-determined idea about what these might be, albeit we must acknowledge that both CMS and JP are medical professionals with clinical experience and academic interest in the management of OHCA. We were guided by our research aim and so sought information about the participants’ interaction with the AED, drone and 999-call-handler.

We aimed to recruit 20 participants. Non-pregnant adults (≥18 years) who felt physically capable of performing CPR in the simulations) were eligible. For reasons of pragmatism this was a convenience sample of (non-healthcare) personnel known to local police, search and rescue operations or members of the study team.

The study received ethical approval from the University of Warwick Biomedical and Scientific Research Ethics Committee (ref: BSREC 109/19-20) on 02/06/2020. Study participants received a Participant Information Leaflet and Consent Form via email at least 48 hours in advance. They had an opportunity on arriving at the simulation (in a separate holding area) to read a hard copy of the Participant Information Leaflet and ask questions, before reading and signing the Consent Form.

We conducted a COVID-safe simulation day in line with government guidelines at the time. Participants performed compression-only CPR on the manikin.

We stored all study data electronically on an encrypted University of Warwick device running Windows 10. We scanned paper-based data collected on study days onto the computer within 48 hours and shredded the originals.

We have reported this article as per STROBE (The Strengthening the Reporting of Observational Studies in Epidemiology) guidelines for observational studies (23), referring to an extension for simulation studies (24).

### Patient and public involvement

MO’S contributed to review and re-draft of project protocol and all participant-facing documents; observed and offered feedback to the study team on study days; reviewed and edited this published article.

## Results

### Developing the delivery system

We conducted five test flights in November 2020. Timings were (seconds):

Landing and detaching AED: 23.6, 21.7, 23.2, 23.3, 22.

Winch and releasing AED: 21.8, 19.4, 19.6, 16.1, 18.6

The winch was slightly faster. The development team inferred this was largely because the winch lowered the AED faster than the drone could descend to the ground. This, and additional concerns about people prematurely approaching a landed drone whose rotors were still operating meant that we proceeded using the winch mechanism.

**Figure 1** shows the drone, AED and its cradle.

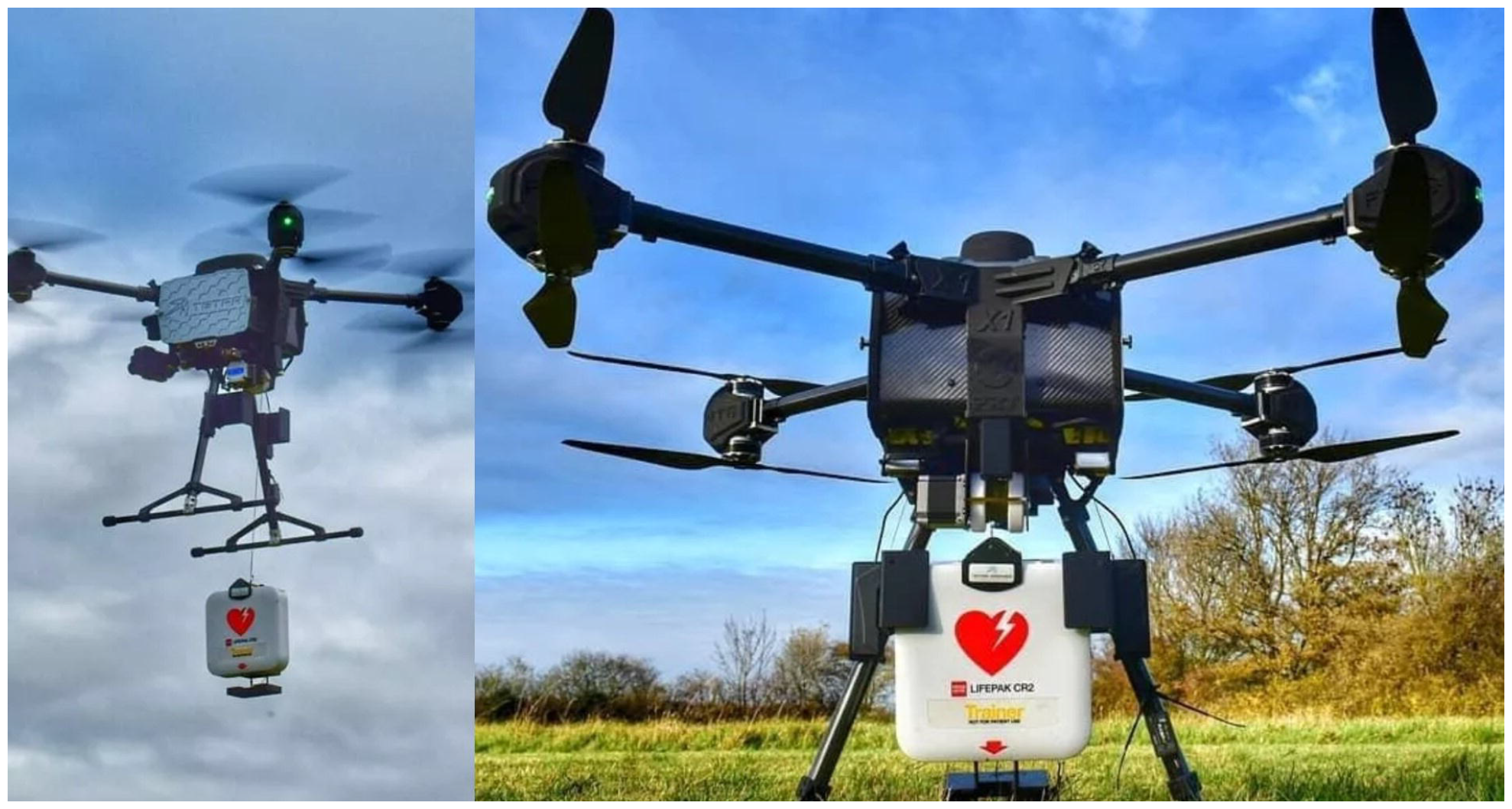

### OHCA simulations

We recruited 18 participants (n=10 female, n=15 previous CPR training); all attended. All participants successfully retrieved the AED, attached it to the patient, delivered a shock and resumed CPR. The median hands-off time was 109s (87-130s). The call-handler answered the 999 call 35s (31-40s) and the drone took off 116s (109-135s) seconds after the simulation started. When retrieving the AED, the participants were away from the patient for 19s (16-22s). **Table 1** has full timings.

**Table 1:** Simulation study timings.

| Simulation component | Time taken (seconds): median (IQR) |
| --- | --- |
| Time to answer 999 call | 35 (31-40) |
| Time to drone taking off | 116 (109-135) |
| Time away from patient's side | 19 (16-22) |
| *Time from patient to reaching AED* | *10 (7-12)* |
| *Time from AED back to patient* | *10 (8-11)* |
| Time from AED arrival to first shock | 103 (81-126) |
| Hands-off CPR time | 109 (87-130) |

The post-simulation questionnaire (**Table 2**) indicated that participants were confident interacting with the drone and did not find it difficult.

**Table 2:** Post-simulation questionnaire.

| Statement | Score: median (IQR) |
| --- | --- |
| "I found it unnecessarily complex" | 1 (1-2) |
| "I thought it was easy to do" | 5 (4-5) |
| "I feel very confident doing this" | 4 (4-5) |
| "I would imagine that most people would be able to do this" | 4 (4-4) |

*Questionnaire scored from 1 (strongly disagree) to 5 (strongly agree)*

Some participants reported uncertainty about whether the device on the ground was the AED they were looking for, although video analyses revealed negligible delays in moving towards the AED in any simulation. The drone noise (arriving and departing) made hearing call-handler and AED instructions difficult. Participants were sometimes uncertain about when it was safe for them to leave the patient to retrieve the AED, even after hearing the siren. Several participants directly asked the call-handler if they could leave the patient before doing so.

Retrieving the AED was easy. However, the cradle at the end of the winch in which it was held (see **Figure 1**) interfered with lying the AED down flat and made it more difficult to access the button required to open the case. There was sometimes conflict between call-handler instructions and AED voice instructions about how to use the AED.

**Table 3** summarises the main themes emerging during post-event interview and audio-visual analyses.

**Table 3:**
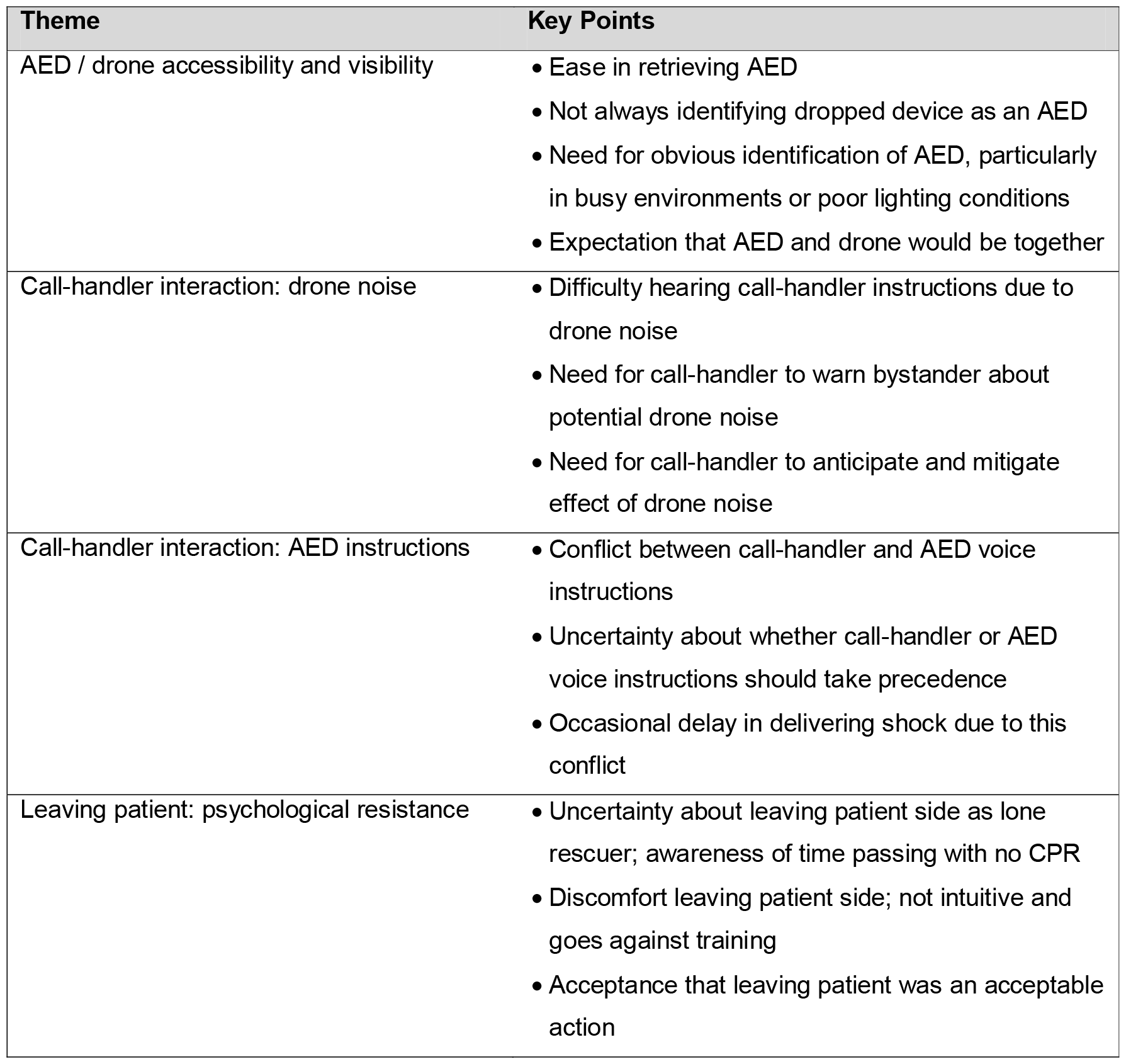
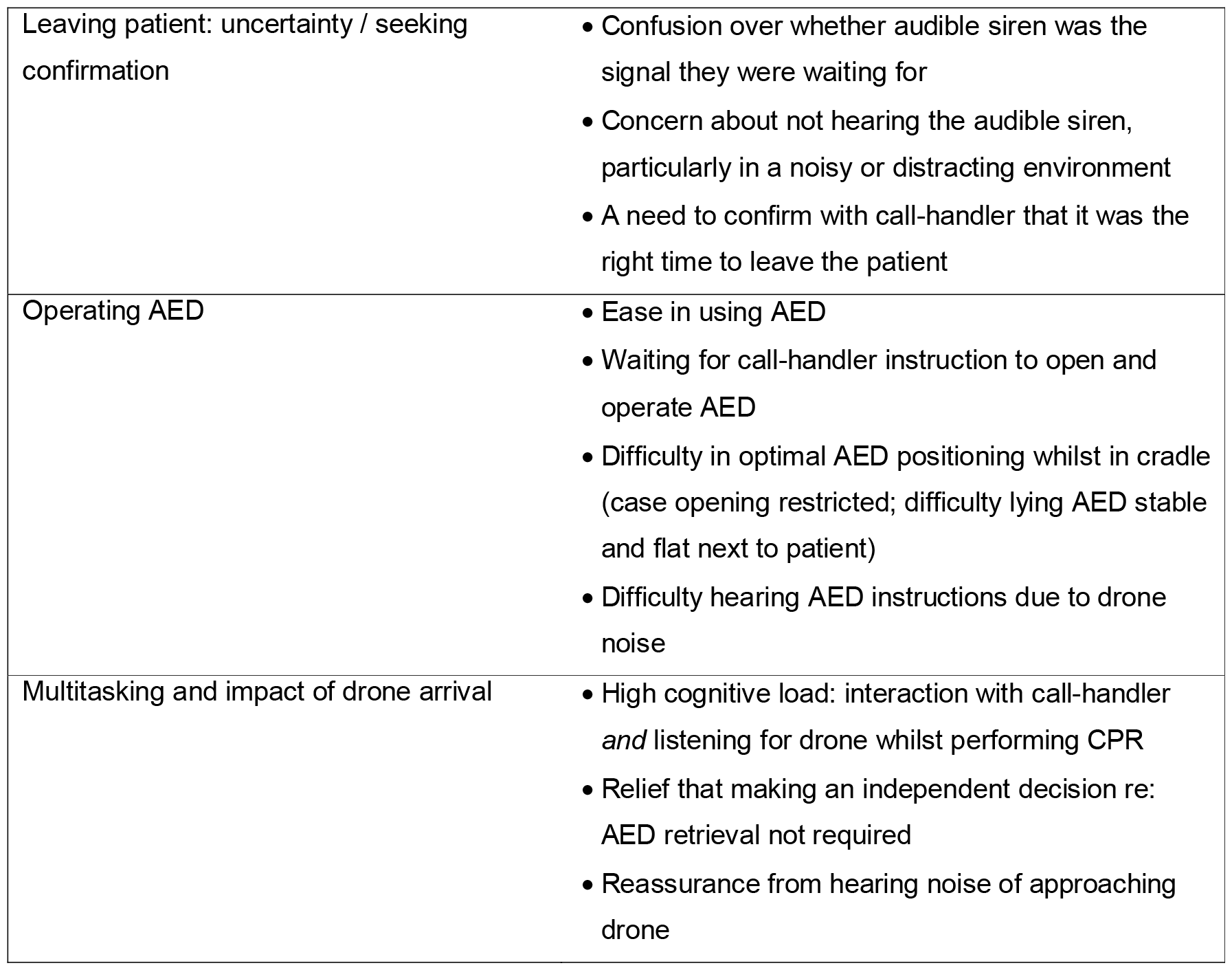
Themes arising from post-event analyses.

A 30s subtitled video of the simulation (mocked-up with JP as the ‘participant’ and CMS’ voice on the phone) is available in the **Electronic Supplementary Material**.

## Discussion

### Main findings

Lowering an AED from a drone via winch was quicker and felt to be safer than the drone landing and the AED detaching. Participants in our lone-rescuer simulations had a median of 109s ‘hands-off CPR’ when retrieving the drone-delivered AED but spent only 19s away from the patient’s side. They found it easy to interact with drone and AED and were willing to do so despite some expressing discomfort at having to leave patient.

Simulations revealed issues about AED visibility and AED stability in its cradle when positioned next to a patient. Drone noise had a substantial impact on participants’ interactions with the call-handler, and there was sometimes conflict between call-handler and AED voice instructions. Whilst the arrival of a drone-delivered AED gave participants an extra thing to consider, some recognised that delivering an AED to their location relieved them of the decision to retrieve one.

### Comparison to the literature

We lowered an AED to the ground via winch using a rotary wing drone. In a previous UK study, researchers used fixed-wing aircraft to carry an AED a total of 92km in six test flights in a coastal air corridor. The AED was dropped by parachute from a height of 120m, and it landed within 50m of the target location (18). This is the first UK study to demonstrate safe AED delivery immediately outside the target location and a subsequent effective interaction between bystander and drone/AED.

We demonstrated a median hands-off CPR time of 109s. In a 2018 simulation study in Sweden, four untrained participants in lone-bystander scenarios retrieved a drone-delivered AED with a hands-off CPR time ranging from 75-110s. That study also reported, like we did, that participants found drone/AED interaction easy, felt relieved when the drone arrived, and perceived that effective communication with the call-handler facilitated AED use (17).

Our study focussed on processes at the end of drone flight, but minimising time delays before flight (i.e. from time of OHCA recognition to the time that the drone is ready to fly) is just as important.

In Germany, researchers performed end-to-end simulations, fully integrating a drone delivery network with local ambulance service systems, for 46 simulated OHCAs across five flight paths. Mean time to defibrillation (from time of emergency call) was 6:02mins for the shortest route (only 0.4km), suggesting substantial delays before and/or after the drone flight itself (25).

AEDs were dispatched via drone for 12 real-world OHCAs in Sweden (June-September 2020), arrived on scene on 11 occasions, and before the ambulance service seven times. However, a drone-delivered AED was not attached to a patient on any occasion. There was a median time delay of 3:10mins from the emergency call starting to the initiation of drone flight: shortening this delay may improve the effectiveness of the system (11).

Drone-delivered defibrillation *is* possible. In the Swedish system (December 2021) a 71-year-old man was defibrillated with a drone-delivered AED and survived to hospital discharge. The drone took off 2:15mins after the emergency call started and the time from emergency call to first shock was 6:15mins (12).

### Strengths and limitations

We have reported on bystander interaction with a drone-delivered AED for the first time in the UK. We examined only the lone-rescuer situation and did not compare this with two-or-more rescuer simulations. We did not consider OHCAs occurring in multi-level buildings or the delay to AED retrieval that this might cause.

We present both quantitative and qualitative data, the latter being useful to explore participants’ experiences and behaviours (26). We ‘triangulated’ data from different sources in an attempt to present a richer picture of the participant experience (27). An understanding of human behaviours is key to the successful implementation of any complex system (28, 29), such as the use of drone-delivered AEDs to complement the community response to OHCA. This is a major strength of the study not explored in this detail before.

Our participant sample was pragmatic and convenient. We did not formally assess data saturation. An appropriate sample size depends on several factors, but will be reduced by the specificity of the situation being investigated (e.g. here, fetching a drone-delivered AED), homogeneity of participants (all performing a standardised simulation), and the briefness of the post-event interview and audio/video clips (30).

Most of our participants had prior CPR training. In the UK (2017), 59% reported previous CPR training and 19% previous AED training, so our group may not be representative of the wider public (31). In the simulations we used a siren to indicate safe drone arrival because effective communication systems between drone and 999 call-handler do not yet exist. Participants indicated that a direct instruction from the call-handler would be a better option than listening for an audible cue to approach the drone.

### Clinical implications

If drone-delivered AEDs are to be successful in real-life OHCAs, researchers must optimise multiple processes. These include preparing an effective network (e.g. modelling optimal drone base locations, deciding on response radius and dispatch rules), and optimising both pre-flight (automated processes to activate drone and pilot following OHCA recognition during a 999 call; liaison with air-traffic control about flight paths) and post-flight (safe AED delivery, with effective bystander interaction) processes (32). Implementing a drone-delivered AED network will have broader implications for the aviation and logistics industries as they seek to perform routine BVLOS drone flight.

In our study, we reported a median hands-off CPR time of 109s. However, participants only spent 19s away from the patient – this is the *additional* hands-off time compared to a lone-rescuer having their own AED on site. We have not seen this latter calculation reported elsewhere. This may be acceptable in a real-world situation. Researchers in an international survey considered that an intervention causing a 5% improvement in OHCA survival was clinically relevant (33). Following collapse, each minute without intervention equates to an approximately 10% decrease in survival (34). We would therefore consider that an addition of 30s or more to safely deliver a shock would be clinically relevant.

Our study demonstrated that the 999 call-handler’s role is vital, and so there is a need for effective real-time communication between the drone, its operator and the call-handler. Future researchers can improve usability and visibility of drone-delivered AEDs to make them even easier to use.

The use of drone-delivered AEDs does not fit into a ‘script’ or protocol (such as AMPDS) used by ambulance services. How and where it should fit in – especially as asking a lone bystander to leave an OHCA patient is not usual practice – is an important topic for future research, as is optimising standardised dispatch-assisted AED instructions to reduce conflict between call-handler and AED voice instructions.

### Next steps

We are working towards the development of a fully integrated drone-delivery system for simulated OHCAs. This will involve automated, near instantaneous, activation of a remote drone system by the ambulance service once a potential OHCA is recognised during a 999 call, beyond-visual-line-of-sight flying (up to 5km) in dedicated flight corridors and real-time flight path mapping. This current study highlighted that good interaction between bystander and call-handler was vital: developing effective real-time communication between drone and ambulance service systems will allow the call-handler to keep the bystander updated about drone progress and instruct them when it is time to retrieve it.

Demonstrating this system will be a pre-requisite to live operations for real-world OHCAs in the UK.

## Conclusion

Participants in simulated OHCA simulations found it easy to interact with and use drone-delivered AEDs. Hands-off CPR time is a concern, but an AED delivered directly to a lone rescuer by drone only slightly increased hands-off CPR time.

## Data Availability

All data produced in the present study are available upon reasonable request to the authors

## Acknowledgements

TETRA drones for their work on developing the drone delivery system, and for flying the drone and providing technical support on simulation days. Steven Prince and colleagues at Sussex Police Training Centre for hosting the simulations and the study team. Christopher Shoebridge for videography on simulation days. The call-handler team at WAST for answering training 999-calls on simulation days.

## Data availability

Simulation study day timings, responses to the questionnaire based on the System Usability Scale, field notes and sorting of these notes into themes are available in the **Electronic Supplementary Material**.

## Conflicts of interest

CMS has volunteer roles with Resuscitation Council UK, European Resuscitation Council and the International Liaison Committee on Resuscitation. He is a National Institute for Health Research-funded Clinical Lecturer in Emergency Medicine.

JP has volunteer roles with Resuscitation Council UK.

NR – no conflict declared

CP – no conflict declared

MO’S – no conflict declared

## References

1. OHCAO Registry. (2021). Out-of-Hospital Cardiac Arrest Overview: England 2020. Available from: https://warwick.ac.uk/fac/sci/med/research/ctu/trials/ohcao/publications/epidemiologyreports/ohca_epidemiological_report_2020_-_england_overview.pdf [last accessed 3rd August 2022].

2. Grasner JT, Wnent J, Herlitz J, Perkins GD, Lefering R, Tjelmeland I, et al. Survival after out-of-hospital cardiac arrest in Europe - Results of the EuReCa TWO study. Resuscitation. 2020;148:218–26.

3. Yan S, Gan Y, Jiang N, Wang R, Chen Y, Luo Z, et al. The global survival rate among adult out-of-hospital cardiac arrest patients who received cardiopulmonary resuscitation: a systematic review and meta-analysis. Crit Care. 2020;24:61.

4. Olasveengen TM, Mancini ME, Perkins GD, Avis S, Brooks S, Castrén M, et al. Adult Basic Life Support: International Consensus on Cardiopulmonary Resuscitation and Emergency Cardiovascular Care Science With Treatment Recommendations. Resuscitation. 2020;156:A35–A79.

5. Holmberg MJ, Vognsen M, Andersen MS, Donnino MW, Andersen LW. Bystander automated external defibrillator use and clinical outcomes after out-of-hospital cardiac arrest: A systematic review and meta-analysis. Resuscitation. 2017;120:77–87.

6. OHCA Steering Group. (March 2017). Resuscitation to Recovery. Available from: https://www.resus.org.uk/library/publications/publication-resuscitation-recovery [last accessed 3rd August 2022].

7. Welsh Government. (June 2017). Out of Hospital Cardiac Arrest Plan. Available from: https://gov.wales/sites/default/files/publications/2019-03/out-of-hospital-cardiac-arrest-plan.pdf [last accessed 3rd August 2022].

8. The Scottish Government. (2015). Out-of-hospital cardiac arrest. A strategy for Scotland. Available from: http://www.gov.scot/Resource/0047/00474154.pdf [last accessed 3rd August 2022].

9. Department of Health Northern Ireland. (2014). Community Resuscitation Strategy Northern Ireland https://www.health-ni.gov.uk/sites/default/files/publications/dhssps/community-resuscitation-strategy-2014.pdf [last accessed 3rd August 2022]

10. Smith CM, Lim Choi Keung SN, Khan MO, Arvanitis TN, Fothergill R, Hartley-Sharpe C, et al. Barriers and facilitators to public access defibrillation in out-of-hospital cardiac arrest: a systematic review. Eur Heart J Qual Care Clin Outcomes. 2017;3:264–73.

11. Schierbeck S, Hollenberg J, Nord A, Svensson L, Nordberg P, Ringh M, et al. Automated external defibrillators delivered by drones to patients with suspected out-of-hospital cardiac arrest. Eur Heart J. 2022;43:1478–1487.

12. Schierbeck S, Svensson L, Claesson A. Use of a Drone-Delivered Automated External Defibrillator in an Out-of-Hospital Cardiac Arrest. N Engl J Med. 2022;386:1953–4.

13. Boutilier J, Brooks S, Janmohamed A, Byers A, Buick J, Zhan C, et al. Optimizing a Drone Network to Deliver Automated External Defibrillators. Circulation. 2017;135:2454–65.

14. Claesson A, Fredman D, Svensson L, Ringh M, Hollenberg J, Nordberg P, et al. Unmanned aerial vehicles (drones) in out-of-hospital-cardiac-arrest. Scand J Trauma Resusc Emerg Med. 2016;24:124.

15. Claesson A, Bäckman A, Ringh M, Svensson L, Nordberg P, Djärv T, et al. Time to Delivery of an Automated External Defibrillator Using a Drone for Simulated Out-of-Hospital Cardiac Arrests vs Emergency Medical Services. JAMA. 2017;317:2332–4.

16. Cheskes S, McLeod SL, Nolan M, Snobelen P, Vaillancourt C, Brooks SC, et al. Improving Access to Automated External Defibrillators in Rural and Remote Settings: A Drone Delivery Feasibility Study. J Am Heart Assoc. 2020;9:e016687.

17. Sanfridsson J, Sparrevik J, Hollenberg J, Nordberg P, Dja□rv T, Ringh M, et al. Drone delivery of an automated external defibrillator – a mixed method simulation study of bystander experience. Scand J Trauma Resusc Emerg Med. 2019;27:40.

18. Rees N, Howitt J, Breyley N, Geoghegan P, Powel C. A simulation study of drone delivery of Automated External Defibrillator (AED) in Out of Hospital Cardiac Arrest (OHCA) in the UK. PLoS One. 2021;16:e0259555.

19. Brooks J. SUS: A ‘Quick and Dirty’ Usability Scale. In: Jordan P, Thomas B, Weerdmeester B, McClelland I, editors. Usability Evaluation in Industry. London: Taylor & Francis; 1996. p. 189–95.

20. Norris E, Bullock I. A ‘Learning conversation’ as a style of feedback. MedEdPublish. 2017;6:42.

21. Braun V, Clarke V. Thematic Analysis. A practical guide. London: SAGE 2022.

22. Mason J. Chapter 8: Organizing and Indexing Qualitative Data. Qualitative Researching 3rd ed. London: SAGE; 2017. p. 187–218.

23. von Elm E, Altman DG, Egger M, Pocock SJ, Gøtzsche PC, Vandenbroucke JP, et al. Strengthening the Reporting of Observational Studies in Epidemiology (STROBE) statement: guidelines for reporting observational studies. BMJ. 2007;335:806–8.

24. Cheng A, Kessler D, Mackinnon R, Chang TP, Nadkarni VM, Hunt EA, et al. Reporting guidelines for health care simulation research: extensions to the CONSORT and STROBE statements. Adv Simul (Lond). 2016;1:25.

25. Carolina Baumgarten M, Röper J, Hahnenkamp K, Thies KC. Drones Delivering Automated External Defibrillators-Integrating Unmanned Aerial Systems into the Chain of Survival: A Simulation Study in Rural Germany. Resuscitation. 2022;172:139–145.

26. Choo EK, Garro AC, Ranney ML, Meisel ZF, Morrow Guthrie K. Qualitative Research in Emergency Care Part I: Research Principles and Common Applications. Academic emergency medicine : official journal of the Society for Acad Emerg Med. 2015;22:1096–102.

27. Mays N, Pope C. Quality in qualitative health research. In: Pope C, Mays N, editors. Qualitative Research in Health Care. 3rd ed. Oxford: Blackwell; 2006. p. 82–101.

28. Michie S, van Stralen MM, West R. The behaviour change wheel: a new method for characterising and designing behaviour change interventions. Implement Sci 2011;6:42.

29. Skivington K, Matthews L, Simpson SA, Craig P, Baird J, Blazeby JM, et al. Framework for the development and evaluation of complex interventions: gap analysis, workshop and consultation-informed update. Health Technol Assess. 2021;25:1–132.

30. Malterud K, Siersma VD, Guassora AD. Sample Size in Qualitative Interview Studies: Guided by Information Power. Qualitative Health Research. 2016;26:1753–60.

31. Hawkes CA, Brown TP, Booth S, Fothergill RT, Siriwardena N, Zakaria S, et al. Attitudes to Cardiopulmonary Resuscitation and Defibrillator Use: A Survey of UK Adults in 2017. J Am Heart Assoc. 2019;8:e008267.

32. Smith CM. Defibrillation for out-of-hospital cardiac arrest. Year of the drone? Resuscitation. 2022;172:146–8.

33. Nichol G, Brown SP, Perkins GD, Kim F, Sterz F, Broeckel Elrod JA, et al. What change in outcomes after cardiac arrest is necessary to change practice? Results of an international survey. Resuscitation. 2016;107:115–20.

34. Valenzuela TD, Roe DJ, Cretin S, Spaite DW, Larsen MP. Estimating effectiveness of cardiac arrest interventions: a logistic regression survival model. Circulation. 1997;96:3308–13.

